# Kinetics of SARS-CoV-2 anti-s IgG after BNT162b2 vaccination

**DOI:** 10.1101/2021.03.03.21252844

**Authors:** Daniel Grupel, Sivan Gazit, Licita Schreiber, Varda Nadler, Tamar Wolf, Rachel Lazar, Lia Supino-Rosin, Galit Perez, Asaf Peretz, Amir Ben Tov, Miri Mizrahi-Reuveni, Gabriel Chodick, Tal Patalon

## Abstract

Deployment of the BNT162b2 mRNA Covid-19 Vaccine in Israel began in December 2020.

This is a retrospective analysis of serological data, describing SARS-CoV-2 anti-S IgG kinetics in 116 Israeli healthcare workers administrated the BNT162b2 vaccine.

Seroconversion occurred by day 14 in all individuals, with IgG levels peaking approximately 30 days post inoculation.

This study demonstrated the kinetics of the antibody response post vaccination with BNT162b2. The robustness of seroconversion was observed, alongside a statistically significant difference in IgG levels between employees over and younger 50 years of afge. Further research is required in order to examine the antibody kinetics overtime, as well as whether the age-dependent difference persists.

## Introduction

Deployment of the BNT162b2 mRNA Covid-19 Vaccine in Israel began on December 19th 2020, following a high efficacy randomized placebo-controlled trial (RCT)^1^.

Recently, analyses based on real world data have been published^2,3^, as different approaches attempt to overcome the difficulty in demonstrating vaccine efficacy in real world non-controlled settings.

The ability to infer immunological protection by serologic tests, as assessed in other infectious conditions, is yet unknown, as data pertaining to antibody levels in relation to clinical outcomes remains limited^4^. This possible correlation (or lack thereof) has significant implications on healthcare policy making.

This descriptive real-world study aims to assess antibody response of vaccinated individuals, by analyzing data collected from healthcare workers in Israel, among the first worldwide to be vaccinated with BNT162b2 in two-dose course 21 days apart. Though analyses of antibody kinetics in response to COVID-19 have been examined^5–7^, antibody kinetics after vaccine administration has yet to be published.

## Methods

### Setting and data collection

Maccabi Healthcare Services (MHS) is the second largest state-mandated, not-for-profit, healthcare provider in Israel with over 2.5 million members. Vaccine rollout in MHS began in December 2020.

As part of the ongoing follow-up and quality control efforts by MHS’ Health Division, serologic tests were made available to its employees in MHS’ central laboratory (MegaLab). Healthcare workers over the age of 18 were serially tested for SARS-CoV-2 specific anti-s antibodies between January 14, 2020 and February 24, 2021 (six weeks).

The serology data was retrospectively analyzed, examining vaccine exposure (calculated as days from the first dose of vaccine administration), age and sex. The study was approved by the MHS’s institutional review board.

### Laboratory methods

SARS-CoV-2 serology testing was performed with Quant II IgG anti-Spike 2-CoV-SARS by Abbott (Illinois, U.S.A.) and reported as AU/mL (Arbitrary units). The cutoff for serology reactivity is 50 AU/mL according to the manufacturer’s instructions.

Values below 21 AU/mL were truncated to 21 AU/mL and similarly for values above 40,000 AU/mL.

### Statistical Analyses

Data were analyzed using the logarithm of the results (means and standard deviations were computed for log (Result)). Mann–Whitney U test was used to ascertain the difference in serological response between age groups using SPSS version 23 (IBM inc.). Two-sided confidence interval for weighted mean of data was calculated using Python. Categorical variables were compared using Chi-squared test or Fisher’s exact test.

## Results

A total of 382 serology tests were performed to 116 MHS’ personnel. For each individual, there were no more than 1 test per week. Baseline characteristics are presented in Table 1.

**Table 1.** Baseline characteristics

| | All<br>(n=116) | Age ≥50<br>(n=68) | Age <50<br>(n=48) |
| --- | --- | --- | --- |
| Age y (mean [±SD]) | 50.3 (11.0) | 57.9 (5.0) | 39.6 (7.6) |
| Sex, male (%) | 9 (7.8%) | 5 (7.4%) | 4 (8.3%) |
| Serology tests, n<br>(mean [±SD]) | 3.3 (1.7) | 3.3 (1.7) | 3.3 (1.7) |
| Days between<br>vaccine doses (mean<br>[±SD]) | 21.5 (1.0) | 21.3 (0.8) | 21.7 (1.1) |

3 individuals had not yet been eligible to receive the second dose of BNT162b2 by the time of data analysis. Two of these are older than 50.

Seroconversion occurred in all individuals in the studied cohort. All samples taken after day 14 were reactive (i.e. - more than 50 AU/mL), no samples were reactive before day 13.

Peak values of anti-s for each participant were observed on day 28.9 (SD 10.7). Individual trajectories of IgG for each subject are presented in Figure 2, while results divided by age groups are presented in Figure 2.

**Figure 1.**
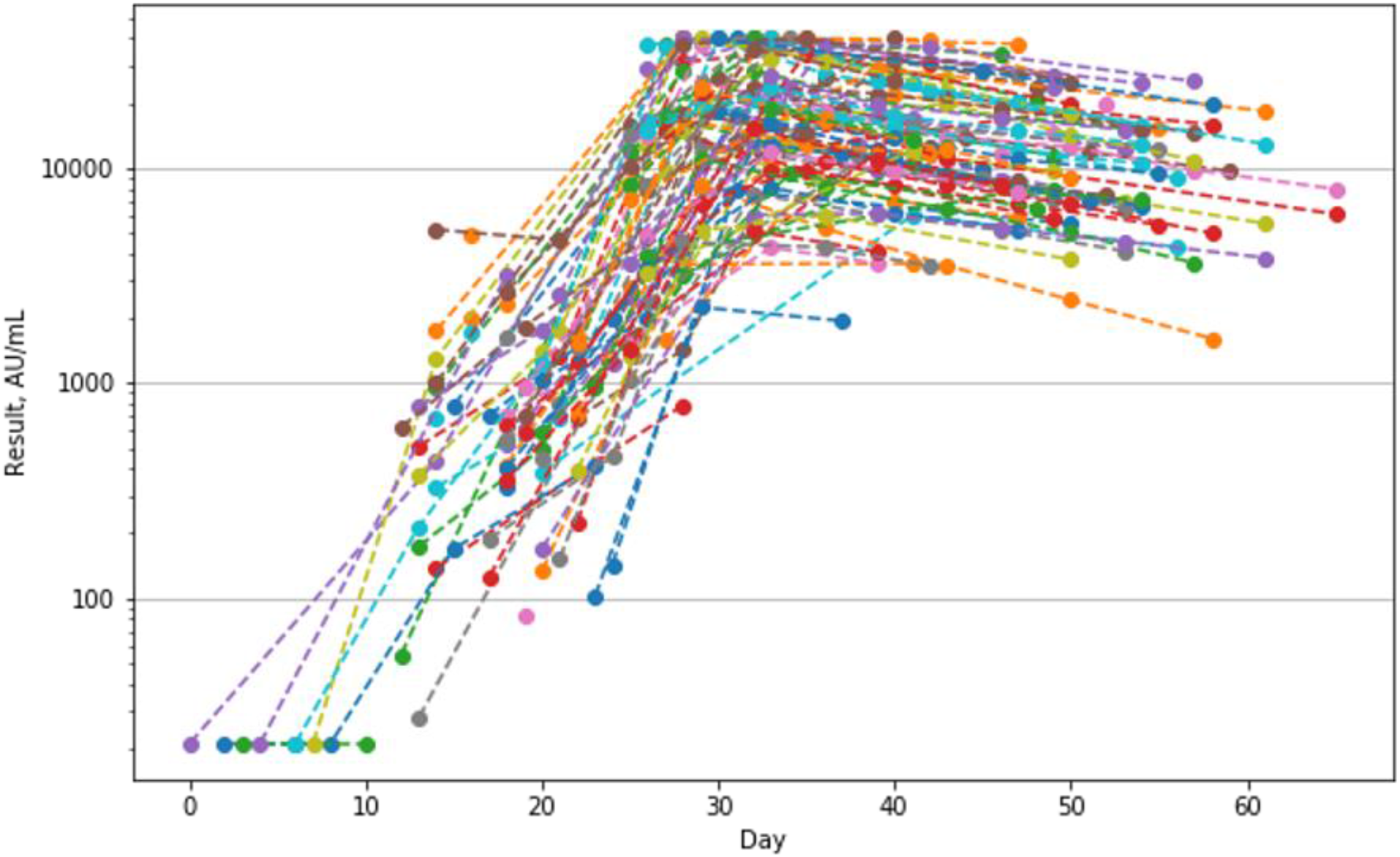
Serology results by days from first vaccine. Each line represents an individual.

**Figure 2.**
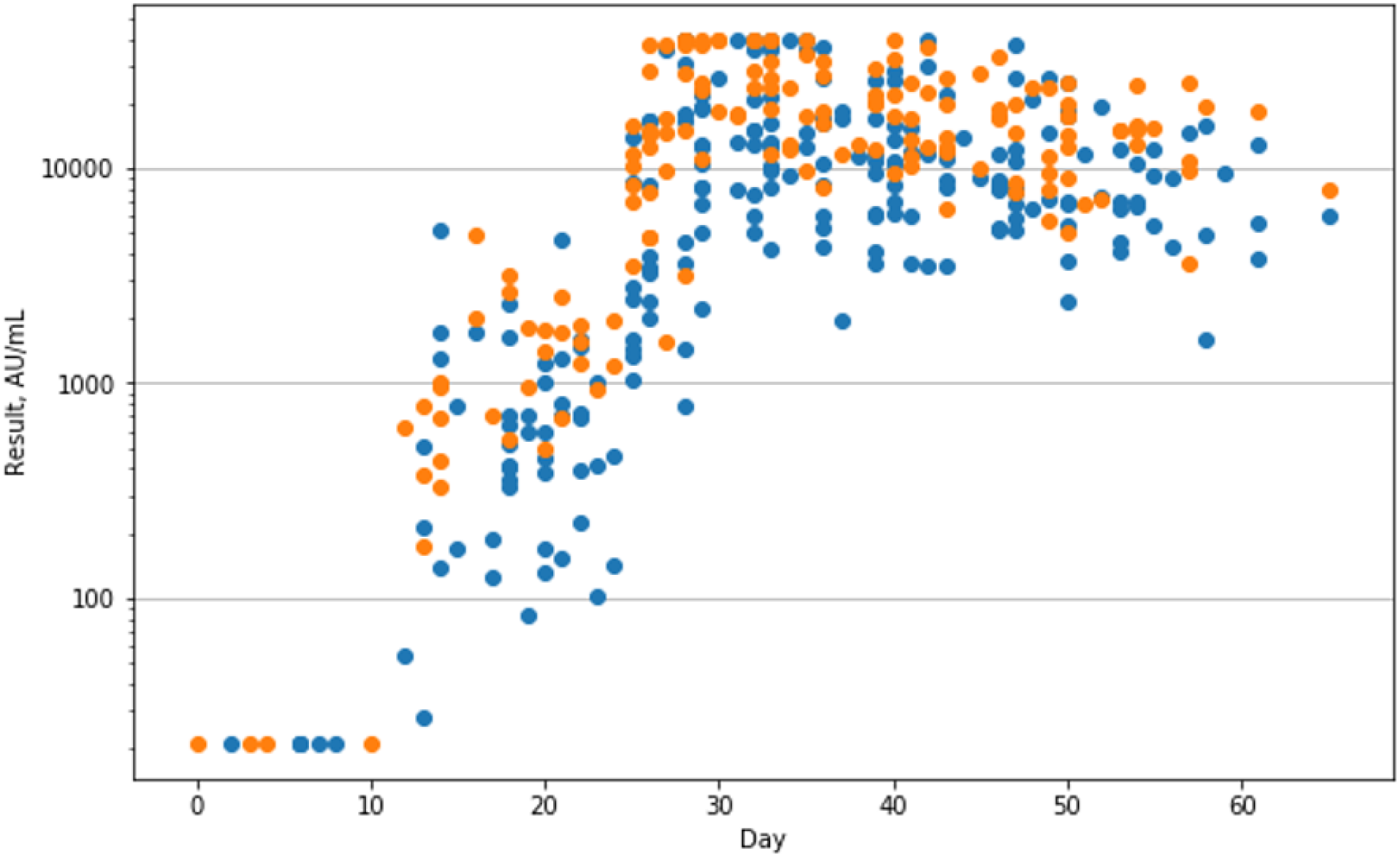
Serology results by days from first vaccine (age ≥50 in blue, age <50 in orange)

Differences were observed in the anti-s IgG levels in members under 50 and over 50. Figure 3 shows serology results per week (with day 0 being the day of the first BNT162b2 dose). Analysis on a per-week basis shows a lasting and statistically significant difference starting at day 15, with the population under 50 having higher IgG levels. It is important to note that while differences do exist, the values are well beyond the cutoff for reactivity.

**Figure 3.**
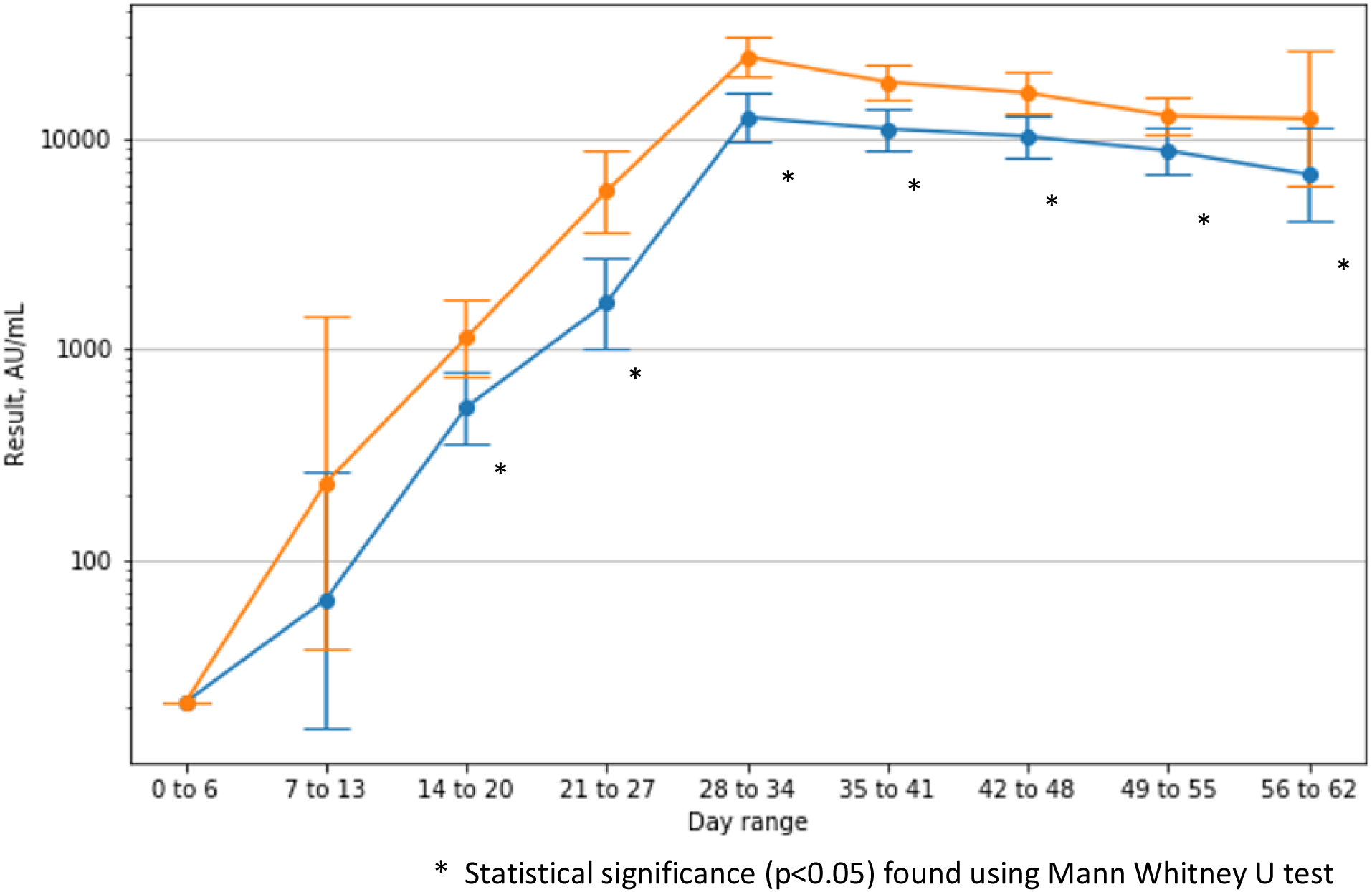
Mean (95% CI) serology result per range of days from first vaccine (age ≥50 in blue, age <50 in orange) * Statistical significance (p<0.05) found using Mann Whitney U test

3 individuals had not yet been eligible to receive the second dose of BNT162b2 by the time of data analysis. Two of these are older than 50.

## Discussion

In our cohort of Israeli healthcare workers, a robust antibody response is demonstrated, starting roughly two weeks after the first dose of BNT162b2 and peaking around day 30 (approximately 10 days after the second dose).

Decline is slow after peaking, though data was limited to six weeks.

A statistically significant difference in IgG levels was observed between employees over the age of 50 and younger individuals, nevertheless, levels remained well above the threshold for serological reactivity.

The importance of this difference needs to be studied further, but a potential difference in vaccine efficacy and vaccine effect length could possibly be present between these two groups.

This study has several limitations. The available cohort is small and not representative of the general population (especially male to female ratio). Moreover, serological test used measures only anti-s IgG.

## Conclusion

This study demonstrated the kinetics of the antibody response post vaccination with BNT162b2. The robustness of seroconversion was observed, alongside a significant age dependent difference in antibody response. Further research is required in order to examine whether the difference persists over time.

More importantly, further investigations should be conducted to evaluate the correlation between seroconversion and clinical course of COVID-19, as well as examining other factors that might affect magnitude of seroconversion as larger datasets become available.

## Data Availability

All data is available with the corresponding author

